# The association of smoking status with hospitalisation for COVID-19 compared with other respiratory viruses a year previous: A case-control study at a single UK National Health Service trust

**DOI:** 10.1101/2020.11.26.20238469

**Authors:** David Simons, Olga Perski, Lion Shahab, Jamie Brown, Robin Bailey

## Abstract

**Background:** It is unclear whether smoking increases the risk of COVID-19 hospitalisation. We examined i) the association of smoking status with hospitalisation for COVID-19 compared with hospitalisation for other respiratory viral infections a year previous; and ii) concordance between smoking status recorded on the electronic health record (EHR) and the contemporaneous medical notes.

**Methods:** This case-control study enrolled adult patients (446 cases and 211 controls) at a single National Health Service trust in London, UK. The outcome variable was type of hospitalisation (COVID-19 vs. another respiratory virus a year previous). The exposure variable was smoking status (never/former/current smoker). Logistic regression analyses adjusted for age, sex, socioeconomic position and comorbidities were performed. The study protocol and analyses were pre-registered in April 2020 on the <u>Open Science Framework</u>.

**Results:** Current smokers had lower odds of being hospitalised with COVID-19 compared with other respiratory viruses a year previous (OR_adj_=0.55, 95% CI=0.31-0.96, *p*=.04). There was no significant association among former smokers (OR_adj_=1.08, 95% CI=0.72-1.65, *p*=.70). Smoking status recorded on the EHR (compared with the contemporaneous medical notes) was incorrectly recorded for 168 (79.6%) controls (χ^2^(3)=256.5, *p*=<0.001) and 60 cases (13.5%) (χ^2^(3)=34.2, *p*=<0.001).

**Conclusions:** In a single UK hospital trust, current smokers had reduced odds of being hospitalised with COVID-19 compared with other respiratory viruses a year previous, although it is unclear whether this association is causal. Targeted post-discharge recording of smoking status may account for the greater EHR- medical notes concordance observed in cases compared with controls.

## 1. Introduction

COVID-19 is a respiratory disease caused by the SARS-CoV-2 virus. There are in excess of 118 million confirmed COVID-19 cases globally, with over 2.6 million deaths reported (Johns Hopkins Coronavirus Resource Center, 2021). Large age and sex differences in case severity and mortality have been observed (Guan et al., 2020), with hypertension, diabetes and obesity identified as important risk factors (Fang et al., 2020). There are *a priori* reasons to believe that current smokers are at increased risk of contracting COVID- 19 and experiencing greater disease severity once infected. SARS-CoV-2 enters epithelial cells through the ACE-2 receptor (Hoffmann et al., 2020). Evidence suggests that gene expression and subsequent ACE-2 receptor levels are elevated in the airway and oral epithelium of current smokers (Brake et al., 2020; Cai, 2020), potentially making smokers vulnerable to contracting SARS-CoV-2. Other studies, however, show that smoking downregulates the ACE-2 receptor (Oakes et al., 2018). In addition, smoking involves repeated hand-to-mouth movements, which may mean smokers are more likely to contract respiratory viruses such as SARS-CoV-2 (Simons et al., 2020). Early data from the ongoing pandemic have not provided clear evidence for an association of smoking status with COVID-19 outcomes, with a living review and unadjusted Bayesian meta-analysis of over 60 studies indicating that current compared with never smokers may be at reduced risk of SARS-CoV-2 infection, while former smokers are at increased risk of hospitalisation, disease severity and in-hospital mortality compared with never smokers (Simons et al., 2021).

Most studies to date have been limited by the lack of appropriate controls, poor recording of smoking status and insufficient adjustment for relevant covariates. Many studies relied on routine electronic health records (EHRs) to obtain data on demographic characteristics, comorbidities and smoking status. This is problematic, as previous research suggests that data on smoking status obtained via EHRs tend to be incomplete or outright inaccurate, with implausible longitudinal changes observed (Polubriaginof et al., 2018). As hospitalised populations differ by age and sex from the general population (Secondary Care Analytical Team, 2020), comparisons of current and former smoking prevalence in hospitalised and non-hospitalised populations are likely biased. There is therefore a need for alternative study designs with relevant comparator groups and adjustment for covariates to better understand the association of smoking status with COVID-19 disease outcomes.

However, the selection of an appropriate comparator group is not straightforward. Ideally, controls should represent the underlying population from which cases emerged, both geographically and demographically (Grimes and Schulz, 2005). In the context of COVID-19 hospitalisation, disease severity and death, we therefore reasoned *a priori* that historical controls – i.e. patients hospitalised at the same trust with another respiratory viral infection (e.g. influenza) a year previous – would act as a useful comparator, as they represent a geographically matched population at risk of severe disease from a circulating respiratory virus with a similar route of transmission (i.e. respiratory droplets and aerosols) and detection (i.e. laboratory-confirmed infection prior to or upon hospitalisation) (McCarthy and Giesecke, 1999). In addition, risk factors for hospitalisation with other respiratory viruses are similar to those for hospitalisation with COVID-19 (e.g. older age, comorbidities) (Falsey et al., 2014; Peralta et al., 2010).

In the present case-control study, we therefore aimed to examine i) the association of smoking status with hospitalisation for COVID-19 compared with hospitalisation for other respiratory viral infections (e.g. influenza, respiratory syncytial virus) a year previous at a single UK hospital trust; and ii) whether there is discordance between smoking status recorded on the summary EHR and within the contemporaneous medical notes. As current smoking in April 2020 (when our study protocol was registered) was *a priori* expected to be associated with an increased risk of COVID-19 hospitalisation (Alqahtani et al., 2020; Simons et al., 2020), and with the association expected to be of a similar magnitude to that observed for other respiratory viruses, we opted for a non-inferiority design to test the hypothesis that the proportion of current smokers in patients hospitalised with COVID-19 is similar to that in patients hospitalised with other respiratory viral infections a year previous.

## 2. Methods

### 2.1. Study design

This was an observational case-control study with historical controls, performed at a single National Health Service (NHS) hospital trust (comprising two hospital sites) in London, UK. The study protocol and analysis plan were pre-registered on the Open Science Framework in April 2020 (https://bit.ly/3kFYh06). The pre- registered protocol stipulated a non-inferiority design (i.e. a one-tailed statistical test) to maximise statistical power to detect a significantly lower proportion of current smokers (i.e. <10%) among patients hospitalised with COVID-19 compared with patients hospitalised with another respiratory viral infection a year previous (i.e. 20%). The protocol was amended after data collection but prior to statistical analysis in September 2020 (https://osf.io/ezfqs/) to implement a traditional case-control design (i.e. a two-tailed statistical test), as a delay in study approval meant that the number of eligible cases and controls exceeded our expectations, providing sufficient power for a two-tailed test. We had also planned to compare current smoking in cases with age- and sex-matched London prevalence, with data obtained from the representative Annual Population Survey. However, following external review on an earlier manuscript draft, we decided against presenting data from this comparison due to smoking rates in hospitalised populations typically being greater than in the general population (Benowitz et al., 2009). A completed STROBE checklist is available in Supplementary file 1. Ethical approval was obtained from a local research committee and approved by the NHS Health Research Authority (IRAS_282704). The requirement for informed consent was waived due to the nature of the study.

A sample size calculation, updated after data collection but prior to data analysis (https://osf.io/ezfqs/), indicated that 363 cases and 109 controls would provide 80% power to detect a 10% difference in current smoking prevalence in cases compared with controls (e.g. 10% in cases and 20% in controls) with alpha set to 5%. We included all cases from 1^st^ March 2020 to the 26^th^ August 2020 (the date on which data were obtained) and all controls from the 1^st^ January 2019 and the 31^st^ December 2019.

### 2.2. Eligibility criteria

#### 2.2.1. Inclusion criteria

##### Cases

1. Consecutive patients admitted to an adult hospital ward (i.e. 18+ years) between 1^st^ March 2020 and 26^th^ August 2020 (the date on which data were obtained);
2. Diagnosis of COVID-19 on or within 5 days of hospital admission, identified via associated International Classification of Diseases version 10 (ICD-10) codes (World Health Organisation, 2019). This temporal boundary was set to prevent inclusion of patents with nosocomial (hospital-acquired) infection and allowed for a delay of 3 days in requesting a COVID-19 test and 2 days for receiving and reporting the results on the EHR. The median incubation time for COVID-19 is estimated at 5.1 days (95% CI = 4.5-5.8) (Lauer et al., 2020). We sought to exclude individuals with nosocomial COVID-19 infection as they are a different population (e.g. older, more frail) compared with those infected in the community and subsequently requiring hospitalisation.

##### Controls

1. Consecutive patients admitted to an adult hospital ward (i.e. 18+ years) between 1^st^ January 2019 and 31^st^ December 2019;
2. Diagnosis of a viral respiratory infection (e.g. influenza, parainfluenza) on or within 5 days of admission, identified via ICD-10 codes.

#### 2.2.2. Exclusion criteria

##### Cases and controls

1. No record of smoking status on the summary EHR or within the medical notes;
2. A primary diagnosis of infectious exacerbation of COPD due to the strong causal association of COPD with current and former smoking.

### 2.3. Measures

Data on demographic and smoking characteristics were collected from the summary EHR or the medical notes. In the UK, the summary EHR is produced at the point of an individual’s first interaction with a specific NHS hospital trust. Further information is added to the summary EHR following subsequent interactions with the hospital trust. The medical notes include contemporaneous clinical notes, General Practitioner referral letters and outpatient clinic letters, and are updated more frequently than the summary EHR.

#### 2.3.1. Outcome variable

The outcome of interest was the type of hospital admission (i.e. with COVID-19 vs. other respiratory viral infections a year previous).

#### 2.3.2. Exposure variable

Smoking status (i.e. current, former, never) was obtained from the summary EHR or the medical notes. A number of cases were recorded as ‘non-smokers’ without distinguishing between ‘former smokers’ and ‘never smokers’. For the primary analysis, patients categorised as a ‘non-smoker’ were treated as ‘never smokers’. Where possible, information on use of smokeless tobacco, waterpipe and/or alternative nicotine products (e.g. e-cigarettes) was extracted. The joint first authors searched within the contemporaneous medical records for free-text entries of smoking status. The most recently available record of smoking status, obtained from either the summary EHR or the medical notes, was extracted. Where available, data on pack-year history of smoking were extracted.

#### 2.3.3. Covariates

Covariates included age, sex, ethnicity, socioeconomic position (SEP; with post codes linked by the research team to the Index of Multiple Deprivation (IMD) (Department for Communities and Local Government, 2019)) and comorbidities (classified by organ system, including cardiac, metabolic and respiratory diseases). Medical conditions not expected to be strongly associated with COVID-19 hospitalisation were not considered in the analyses (e.g. sciatica and fibromyalgia; see Supplementary file 1). Age was treated as a continuous variable in the primary analysis, with banded age groups (i.e. 18-29 years, 30-44 years, 45-59 years, 60-74 years, 75-89 years and > 90 years) used in exploratory analyses. The IMD was categorised as quintiles to reduce the impact of sparse data.

### 2.4. Data analysis

All analyses were conducted in R version 4.0.2.(R Core Team, 2020) Descriptive statistics for cases and controls are reported. To explore differences between cases and controls, Pearson’s Chi-square tests, Cochran-Armitage tests for trend and ANOVAs were used, as appropriate.

To examine the association of former and current smoking with hospitalisation for COVID-19 compared with hospitalisation for other respiratory viral infections, unadjusted and two different adjusted generalised linear models with a binomial distribution and logit link function were performed. The first model adjusted for age, sex and SEP, with a second model adjusting for age, sex, SEP and comorbidities. We report odds ratios (ORs), 95% confidence intervals (CIs) and *p-*values. Two sensitivity analyses were subsequently performed. First, those recorded as ‘non-smokers’ were removed from the analysis. Second, those excluded from the analytic sample due to missing data on smoking status (see section above on ‘Exclusion criteria’) were included and coded as i) ‘never smokers’ and then as ii) ‘current smokers’ to assess the robustness of the associations.

To examine the concordance between smoking status recorded on the summary EHR and within the contemporaneous medical notes, Pearson’s Chi-squared tests were performed for the entire sample, and then separately for cases and controls.

## 3. Results

A total of 610 potential cases and 514 potential controls were identified. A total of 446 cases and 211 controls were included in the analytic sample (see Figure 1). Thirteen potential controls and 60 potential cases were excluded due to not having a record of documented smoking status. This was likely due to patients having no prior contact with the NHS foundation trust. Notably, 37 (62%) of potential cases that were excluded because of missing smoking status did not survive to hospital discharge, with no in-hospital mortality in potential controls, which suggests that data may be missing due to increased mortality in cases.

**Figure 1.**
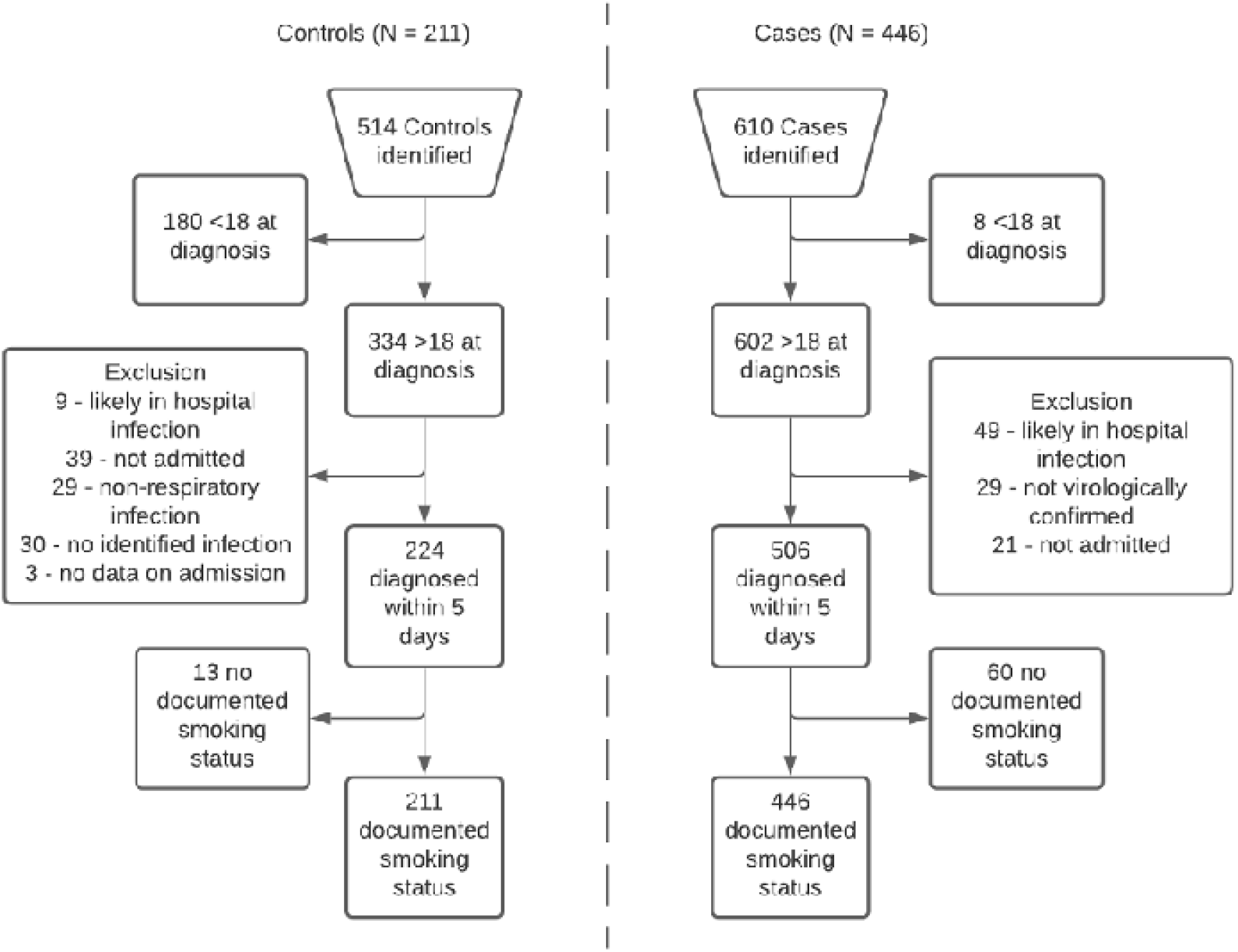
Eligibility flow diagram for controls (left hand side) and cases (right hand side).

Compared with controls, cases were more likely to be male (55% vs. 35.9%) and older (64.9 years vs 62.5 years) (see Table 1). Approximately 10% of cases and controls had missing data for ethnicity. Compared with cases, controls were more likely to be admitted from more deprived areas (IMD quintiles 1 and 2) (41.8% vs. 32.9%, *p* < 0.001). Cases were more likely than controls to have pre-existing metabolic (30.3% vs 13.3%) and cardiac comorbidities (53.4% vs 30.3%). A significantly larger proportion of cases compared with controls did not survive to discharge (28.7% vs. 4.3%). Among 128 cases not surviving to discharge, 53 (41.4%) were never smokers, 63 (49.2%) were former smokers and 12 (9.4%) were current smokers. For patients who survived to discharge, the median length of hospital stay for cases and controls was 9 (IQR = 4-18) and 4 (IQR = 2-9) days, respectively (see Table 1).

**Table 1.**
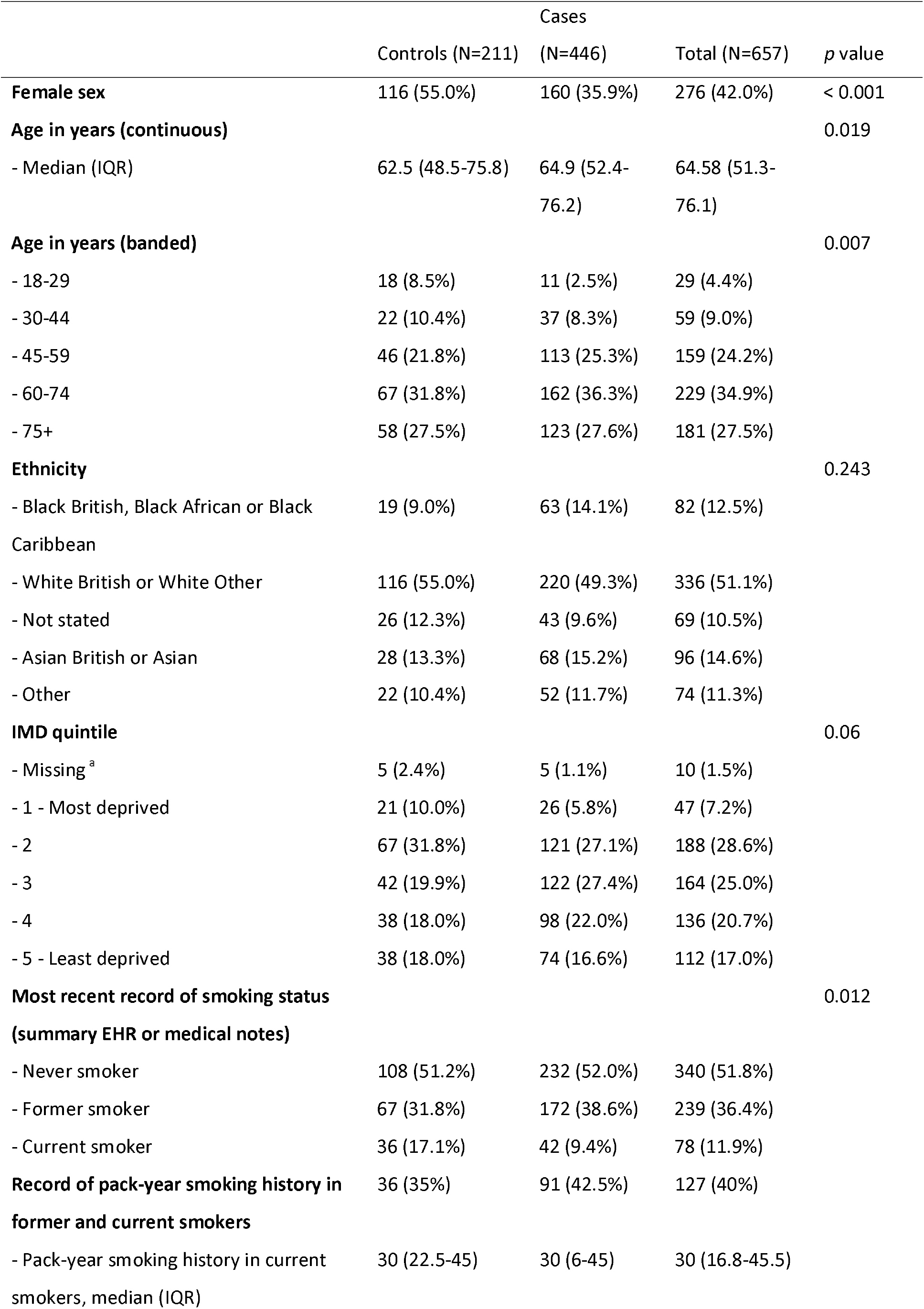
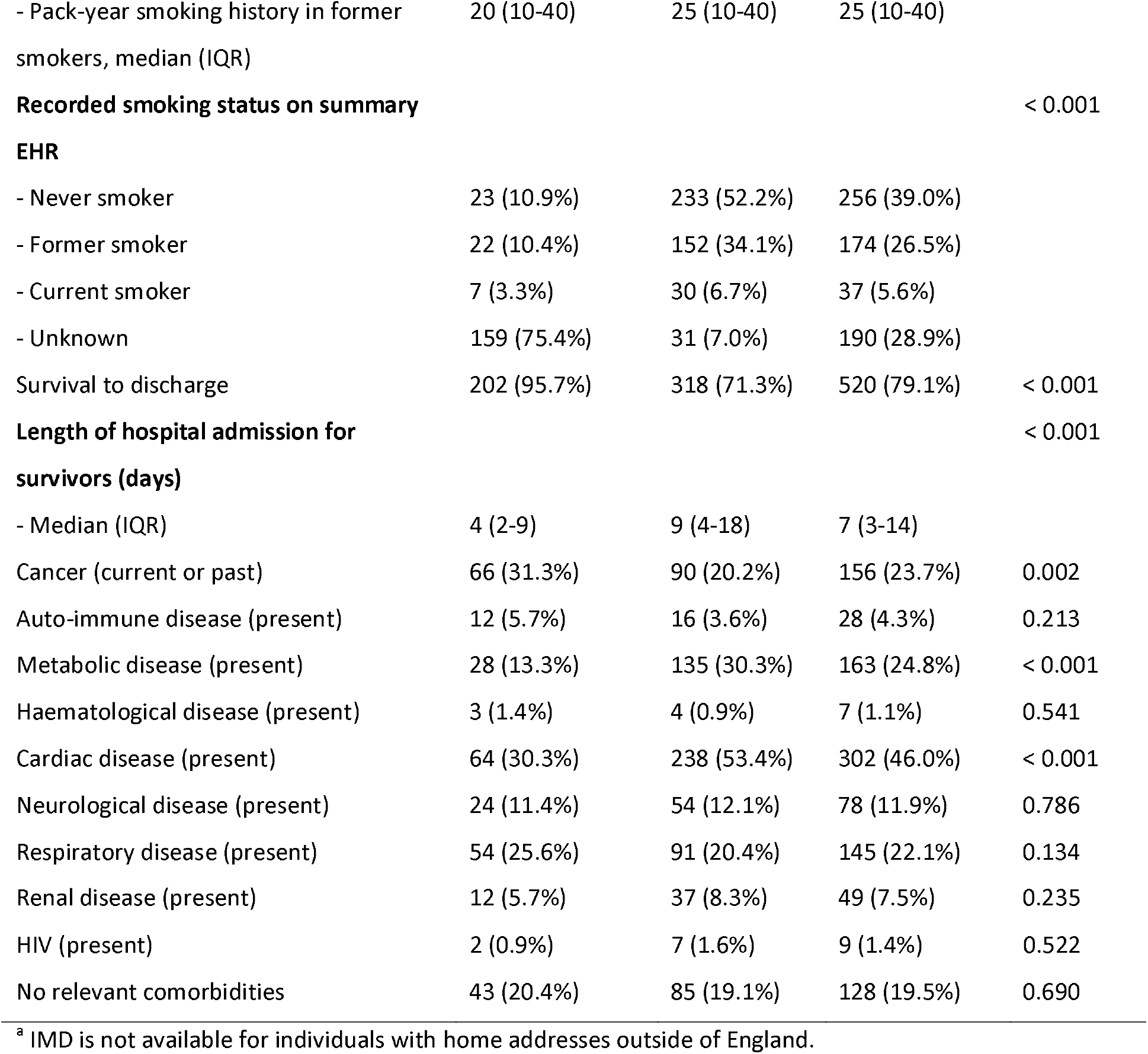
Demographic and smoking characteristics of cases and controls.

Cases and controls were predominantly admitted from North central and North East central London (see Supplementary file 1). The number of cases admitted from peripheral locations was greater than in controls and represents transfer of inpatients from other hospitals and diversion of patients that would otherwise have attended local hospitals due to bed pressures. The Chi-square test for trend found inconclusive evidence for any difference in SEP between cases and controls, χ^2^(3) = 8.93, *p* = 0.06 (see Supplementary file 1).

### 3.1. Association of smoking status with type of hospitalisation

The prevalence of former smoking was higher in cases compared with controls (38.6% vs. 31.8%). Current smoking prevalence was lower in cases compared with controls (9.4% vs. 17.1%). A single patient from the case cohort was recorded as a dual cigarette and e-cigarette user. Two patients, one from each cohort, were recorded as dual cigarette and shisha/waterpipe users. Pack-year history of smoking was only recorded for 40% of patients with a smoking history (see Table 1).

In the univariable analysis, current smokers had reduced odds of being hospitalised with COVID-19 compared with other respiratory viruses a year previous (OR = 0.52, 95% CI = 0.31-0.86, *p* = 0.01). The odds for former smokers were equivocal (OR = 1.16, 95% CI = 0.81-1.68, *p* = 0.43).

In the multivariable analysis adjusted for sex, age and SEP, current smokers had reduced odds of being hospitalised with COVID-19 compared with other respiratory viruses a year previous (OR = 0.48, 95% CI = 0.28-0.83, *p* < 0.01). There was no significant association among former smokers (OR = 0.90, 95% CI = 0.61- 1.34, *p* = 0.61). Results were not materially altered when also adjusting for relevant comorbidities (current smokers, OR = 0.55, 95% CI = 0.31-0.96, *p* = 0.04; former smokers, OR = 1.08, 95% CI = 0.72-1.61, *p* = 0.70).

### 3.2. Sensitivity analyses

First, in a sensitivity analysis with patients recorded as ‘non-smokers’ excluded from the sample (leaving 398 cases and 159 controls), current smokers had reduced odds of being hospitalised with COVID-19 compared with other respiratory viruses a year previous (OR = 0.41, 95% CI = 0.22-0.74, *p* = 0.03). There was no significant association among former smokers (OR = 0.78, 95% C.I. = 0.49-1.23, *p* = 0.28).

Second, in a sensitivity analysis with those with missing data on smoking status (n = 73) treated as ‘never smokers’ (resulting in 506 cases and 224 controls), patients hospitalised with COVID-19 had reduced odds of being a current smoker compared with those admitted with other respiratory viruses (OR = 0.51, 95% CI = 0.30-0.88, *p* = 0.01). Next, when those with missing data on smoking status were treated as ‘current smokers’, there was no significant association between current smoking and hospitalisation with COVID-19 (OR = 0.94, 95% CI = 0.61-1.46, *p* = 0.80).

### 3.3. Concordance of smoking status recorded on the summary EHR and the medical notes

Controls were more likely to have no record of smoking status on the summary EHR compared with cases (75.4% vs. 7%) (see Figure 2). However, smoking status could be ascertained from the contemporaneous medical notes for all included cases and controls. Smoking status on the summary EHR (including ‘unknown’ status) was incorrectly recorded for 168 (79.6%) controls and 60 cases (13.5%) (χ^2^ (3) = 226.7, *p* = < 0.001). In cases, six current smokers were misclassified as former smokers, one current smoker as a never smoker and six current smokers had no record of smoking status on the summary EHR. In controls, six current smokers were misclassified as former smokers and 23 current smokers had no record of smoking status on the summary EHR. There was greater discordance between smoking status recorded on the summary EHR and within the contemporaneous medical notes in controls (χ^2^ (3) = 256.5, *p* = < 0.001) than in cases (χ ^2^(3) = 34.2, *p* = < 0.001).

**Figure 2.**
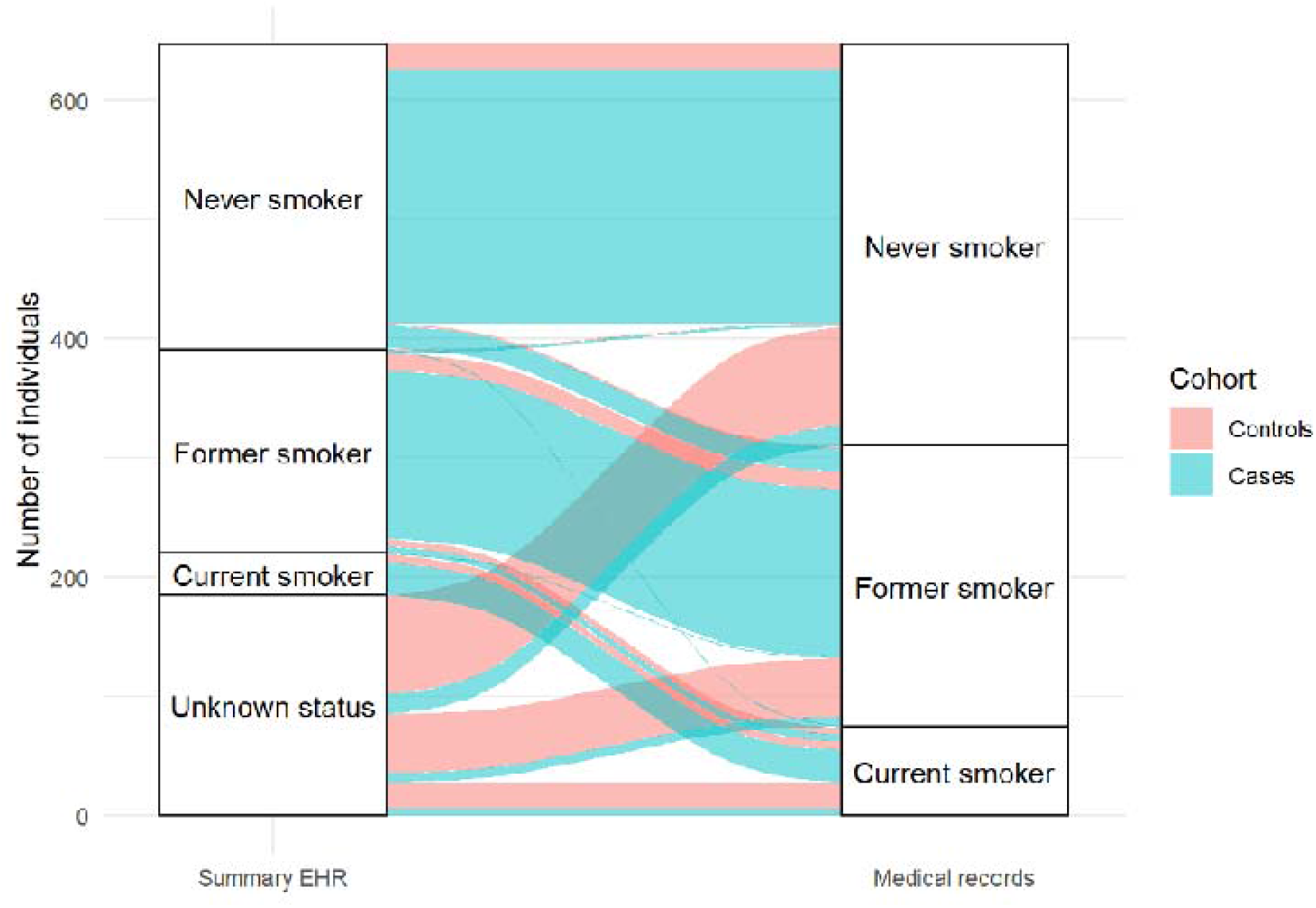
Concordance between smoking status recorded on the summary EHR and the medical notes for controls (red) and cases (blue).

## 4. Discussion

This observational case-control study with patients admitted to a single UK hospital trust found a lower proportion of current smokers in cases hospitalised with COVID-19 during the first phase of the pandemic compared with controls hospitalised with other respiratory viral infections a year previous. Further, we found that smoking status is typically poorly recorded in the summary EHR. This was more prominent in controls than cases – a difference that is likely explained by the observation that COVID-19 patients were followed up by the respiratory medicine team after discharge as part of a COVID-19 follow-up clinic where they specifically asked about smoking status (Mandal et al., 2020). The observed discrepancy between smoking status recorded on summary EHRs and the contemporaneous medical notes is a concern, particularly for studies relying solely on EHRs as the source of information on smoking status.

### 4.1. Strengths and limitations

To our knowledge, this is one of few studies specifically designed to examine the association between smoking status and hospitalisation with COVID-19. It was further strengthened by an assessment of the quality of data on smoking status gleaned from summary EHRs.

However, this study has several important limitations, the majority of which pertain to the selection of the controls. First, current smoking is expected *a priori* to be associated with hospitalisation for non-COVID-19 respiratory viruses (Stämpfli and Anderson, 2009). Ideally, hospital-based case-control studies should avoid selecting a control disease which is associated with the exposure of interest (i.e. smoking status) (Vandenbroucke and Pearce, 2012). However, to our knowledge, there is no other control disease with a similar route of acquisition and mechanism for hospitalisation/severe disease that is not *a priori* also associated with smoking status. The greater smoking prevalence in controls compared with the general population from which the cases emerged (Vandenbroucke and Pearce, 2012) therefore likely contributes to the significantly reduced odds of current smoking in our cases.

Second, the risk profile for controls likely differs from cases in that there is prior immunity to other respiratory viruses (e.g. influenza, respiratory syncytial virus), with no prior immunity in the population to SARS-CoV-2.

Third, we selected the controls on the basis of sharing a similar route of transmission and risk factors for hospitalisation as cases. However, at the time of writing (March 2021), we now suspect that COVID-19 differs from other respiratory viruses in several ways. For example, COVID-19 gains cell entry via the ACE-2 receptor (Hoffmann et al., 2020) – with unknown receptor binding in flu (Killingley and Nguyen-Van-Tam, 2013) – and appears to display less fomite and physical contact transmission than flu (Ben-Shmuel et al., 2020). In addition, emerging evidence suggests that COVID-19 has a significantly different pathological process compared with other respiratory viruses. For example, mortality rates from COVID-19 differ widely from those due to epidemic influenza (Office for National Statistics, 2020a). Although we currently do not know the importance of these factors, taken together, emerging observations may mean that direct comparison of risk profiles in cases and controls is limited.

Fourth, while no known behavioural restrictions were implemented during the control period, London was under lockdown restrictions from March to July 2020, which likely impacted the risk of viral exposure in cases (Davies et al., 2020). This may further have impacted the different risk profiles of controls and cases beyond the adjustments made in this analysis for sex, age and SEP.

Fifth, the selection of historical controls may mean that there are non-trivial differences in smoking status between controls and cases due to a declining trend in London smoking prevalence (Office for National Statistics, 2020b). However, as a single year was used for the selection of controls, and there was no large change in national smoking prevalence from 2019 to 2020 in England (Brown, 2020), we expect any impact of the time-varying exposure to be minimal. We considered using a contemporaneous control (i.e. patients hospitalised with other respiratory viral infections in 2020), which would have mitigated against this potential bias. However, due to factors such as reduced national and international travel, physical distancing, increased hand hygiene and potential viral dominance by COVID-19, exposure to and hospitalisation with other respiratory viruses has been substantially reduced in 2020 (GOV.UK, 2020), which would have limited the sample size for controls.

Sixth, a history of current or past cancer was high in both groups at greater than 20% and was significantly greater in controls compared with cases. This reflects a bias in the population that regularly interacts with the selected NHS hospital trust, which is a specialist cancer referral centre. We visualised the geographic regions where patients were admitted from to examine any systemic differences between cases and controls, and caution that the differing catchment areas of the two cohorts may have led to important differences in the underlying populations. In addition, during the peak of the first wave of the pandemic in the UK, many cases were transferred across hospital sites due to bed pressures.

Finally, there was a greater proportion of cases than controls with no record of smoking status on either the summary EHR or within the contemporaneous medical notes (thus, they were not eligible for inclusion), with patients with missing data having unusually high mortality. It is plausible that many of them were smokers. We attempted to mitigate against this by conducting several sensitivity analyses, with the results largely remaining robust. However, when those excluded from the cohort due to missing data on smoking status were treated as current smokers, there was no significant association between current smoking and hospitalisation for COVID-19.

Despite these limitations, alternative designs were impracticable or would have had different limitations. In the future, the current study can be considered alongside findings across multiple such alternative methodological approaches, each with different sources of bias, to triangulate on the extent to which associations between smoking and COVID-19 are causal.

### 4.2. Implications for policy and practice

COVID-19 will continue to place a large burden on healthcare services in the UK and internationally over the coming months and years. To mitigate against this, multiple non-pharmacological interventions are being implemented to reduce the intensity of demand on acute and intensive services. Irrespective of any direct link between smoking and COVID-19 disease outcomes, smoking is a significant cause for healthcare demand globally. We have argued elsewhere for the need to ramp up smoking cessation support to reduce the current and future burden on healthcare and social services (Simons et al., 2020).

### 4.3. Avenues for future research

The selection of appropriate controls in hospital-based case-control studies is very challenging for a novel respiratory virus such as COVID-19 (which means we converged on a hybrid approach, combining elements from hospital-based case series and case-control designs with historical controls). We recommend the use of representative population-studies with data from multiple sites and with purposeful acquisition of smoking status to better understand the role of smoking as a potential risk or protective factor for COVID- 19 hospitalisation and disease severity.

## 5. Conclusion

In a single hospital trust in the UK, current smokers had reduced odds of being hospitalised with COVID-19 compared with other respiratory viruses a year previous, although we caution against interpreting this as a causal association. Smoking status was poorly recorded, with high observed discordance between smoking status recorded on the summary EHR and the contemporaneous medical notes.

### Data sharing

Anonymised and de-identified individual-level data are available upon request and approval from the NHS foundation trust. Requests should be directed to the corresponding author. Alternatively, data can be requested directly from the Biomedical Research Centre Clinical and Research Informatics Unit at UCL/UCLH.

## Contributors

DS, OP, LS, JB and RB conceived of and designed the study. DS and RB acquired the study data. DS and OP extracted and analysed the data. DS, OP, LS, JB contributed to the interpretation of the data and to the drafting of the manuscript. All authors have read and approved the manuscript.

## Acknowledgements

DS is supported by a PhD studentship from the UK Biotechnology and Biological Sciences Research Council [BB/M009513/1]. OP receives salary support from Cancer Research UK [C1417/A22962]. JB, LS, & OP are members of SPECTRUM, a UK Prevention Research Partnership Consortium [MR/S037519/1]. UKPRP is an initiative funded by the UK Research and Innovation Councils, the Department of Health and Social Care (England) and the UK devolved administrations, and leading health research charities.

## Ethical approval

This study was approved by the UCL/UCLH Joint Research Office Research Strategy Group and UCLH Data Access Committee. Approval to conduct research limited to pseudonymised patient data was provided by the NHS Health Research Authority (IRAS_282704). The requirement for informed consent was waived due to the nature of the study.

